# Gender differences in correlations of self-reported marijuana use with a creatinine-normalized urine biomarker among persons with HIV

**DOI:** 10.1101/2025.03.14.25323995

**Authors:** Donald D. Porchia, Yancheng Li, Michael Truver, Bruce Goldberger, Eric C. Porges, Yan Wang, Zhigang Li, Samuel Wu, Robert L. Cook

## Abstract

**Background:** Cannabis use is common among persons with HIV (PWH). However, accurately quantifying cannabis consumption and its components such as THC has been a long-standing challenge due to the variety of cannabis products available, variability in smoking and other consumption behaviors, as well as the reliability and validity of self-reported cannabis measures. Further, there is a paucity of research on the relationship between a timeline follow back (TLFB) and cannabis metabolites/biomarkers in older, underrepresented populations of PWH. We sought to determine the association between the self-reported quantity of cannabis used with the creatinine-normalized urine biomarker (THC-COOH) primarily among exclusive flower users and as a secondary analysis among cannabis users of any method of administration. We also explored whether the associated effect size differed by gender at birth.

**Method:** Data were collected from 2018-2022 from participants in the MAPLE study, a longitudinal study examining health effects of cannabis among PWH in Florida. This analysis included data from 253 PWH (mean age 49, 41% Female, 16% White, 68% Black, and 13% Hispanic) who used cannabis products. The quantity of daily cannabis use over the past 30 days was collected by trained interviewers at three time points (baseline, 1 year and 2 years follow-ups) using a self-reported, calendar-based TLFB. Using these findings, cannabis quantity was summarized in terms of mg of THC per day and urine THC-COOH, a biomarker for THC, was determined by mass spectroscopy from 103 participants. A linear mixed effect model was used to test the association between the creatinine-normalized THC-COOH levels and average quantity of mg THC consumed per day for exclusive marijuana flower users and also for all users of any marijuana product. We also stratified by gender to investigate the effect size in males and females.

**Results:** Among the exclusive marijuana flower users we found an association between mg of THC per day and THC-COOH urine biomarkers, overall and in both males and females. The strength of the fixed effect was stronger in females than males. However, when we examined users of any marijuana product, not just flower, we did not find a significant association between mg of THC per day and THC-COOH urine biomarkers among males or females.

**Conclusions:** TLFB is a valid instrument to measure self-reported marijuana flower use. However, the fixed effect between self-reported consumption of cannabis with urine THC-COOH biomarkers is stronger in females than in males.

## 1. Introduction

Cannabis use is widespread and reported to be increasing^1,2^, with an estimated 19.8% of the North American population reporting use in 2022^3^. With such widespread use, it is important to understand the impact of cannabis components such as delta9-THC on health. Cannabis is commonly used to help manage a variety of symptoms such as chronic pain or nausea and various mental health conditions such as depression, anxiety and post-traumatic stress disorder, as well as recreationally. In order to better understand the consequences of marijuana use, particularly in the context of dose response relationships, accurate and low burden cannabis consumption metrics are vital for researchers investigating such health outcomes.

Yet measuring cannabis use is inherently difficult. There are variety of methods of methods of administrations such as inhalation through joints, blunts, bongs or vaporizers. It may also be ingested through edibles in the form of gummies, brownies or other consumables and the types of products are only increasing with increased legalization.

In order to calculate the amount of THC ingested we need to know the method of administration. For smoked flower one must estimate the percentage of THC in the flower. Yet when purchasing flower from medical marijuana dispensaries there is reason to question the accuracy of the THC labelling, One study found that as much as 70% of samples of THC levels from dispensary cannabis were more than 15% lower than labeled and another study showed it could be even higher than labelled^4,5^. Additionally, the THC levels of the majority of cannabis obtained from outside the medical marijuana system is unknown. This makes converting from quantity of grams smoked to quantity of THC ingested problematic. There are similar issues when considering edibles either obtained from dispensaries or “cooked” at home. Further, although blood and urine toxicology can be accurate, it is prohibitively expensive for routine use. Whereas, individual self-reports are both easily measurable and cost effective, but they have failed to provide an accurate measure of cannabis use^6–11^.

We proposed using a timeline follow back (TLFB) to calculate the average mg of THC ingested at each study visit. We then conducted a longitudinal analysis to examine the association between the self-reported average mg of THC ingested as calculated by the TLFB and the urine THC-COOH, a biomarker for THC from urine samples collected for both the exclusive flower users and users of any type of cannabis product. Further, we examined how the association differed by gender in each case.

## 2. Methods

### 2.1 Study Design

This analysis used data from marijuana users with HIV enrolled in the Marijuana Associated Planning and Long-term Effects (MAPLE) study. Full details of the study including overall study aims, participants and data collection procedures can be found in study protocol^12^ and elsewhere^13^. All research procedures were approved by the Institutional Review Boards at the University of Florida, Florida International University, and the FDOH. All participants provided informed consent before they participated in the study.

### 2.2 Participants

A total of 333 participants were enrolled in the MAPLE study, 260 of which were marijuana users and 73 were control participants who did not use marijuana. Of the 260 marijuana users, 253 were used in the final analysis of which 208 were exclusively flower users and 45 used flower along with other cannabis products. Of the 103 participants whose urine samples were analyzed for THC-COOH urine biomarkers, non-users were removed from the analysis so as not to zero-inflate the data, leaving 81 participants of which 41 were males and 40 were females.

### 2.2 Time line follow back (TLFB)

A Timeline Follow back is a research instrument used to help participants estimate the frequency and quantity of different substances used over a given time period. In this study, a retrospective, calendar-based TLFB was used that had been initially developed for alcohol use but modified to measure cannabis consumption. It had been previously found reliable for use with cannabis and other substances, such as cocaine and tabacco^14–16^. Trained MAPLE research assistants used this TLFB to assess patterns of use related to the timing of daily doses, the method of consumption (e.g. joints, blunts, bowls, etc.), the quantity of marijuana flower consumed in grams, the dosages of other cannabis products consumed, and frequency of use over the past 30 days since each study visit in order to aid in recall^13^.

### 2.3 Outcome variable

The outcome variable used was creatine-normalized THC-OOH concentration in urine. THC-COOH was quantified by isotope-dilution gas chromatography mass spectrometry following solvent extraction from urine. Urine creatine was calculated using Cayman’s kit.

We used creatinine-normalized THC-COOH concentrations because they reduce variability in the concentration of THC-COOH due to hydration effects^17^. From the total population of marijuana users and non-users, 103 participants were selected to provide a diversity of outcomes.

### 2.4 Covariate variables

The TLFB recorded the daily cannabis usage from the previous 30 days at each study visit: baseline, one year follow-up and two year follow-up. Dosages of flower were measured in grams per day while dosages of other cannabis products were measured in mg of THC. See table 1 for the amount of mg of THC assumed to be consumed for each dose by product.

**Table 1.** THC per dosage of Non-Flower Cannabis Products.

| Non-Flower Cannabis Product | Mg of THC |
| --- | --- |
| THC oil vaped | 4 mg per hit/puff/dose if given otherwise 10 mg per day |
| Edible (chocolate or butter) | 20 mg per dose |
| Edible (gummy) | 10 mg per dose |
| Marinol pill | 2.5 mg per pill |
| Dab/Concentrate | 25 mg per day |

For the flower product, if the percentage was known then that percentage was used to convert the grams per day into mg of THC per day. However, since most of the flower was from non-dispensary sources the percentage of THC in the flower was largely unknown. In this case, we examined the cannabis potency data from the National Institute on Drug Abuse (NIDA), which measured the average THC content of cannabis confiscated by law enforcement from 1995 – 2022^18^. From the data, once can see that the THC content has a strong linear correlation over time, raising from an average of approximately 4% THC in 1995 to approximately 16% in 2022. Therefore, in the cases where the THC content was unknown, we choose to use assume an average of 15% THC. For each exclusively flower using participant, we determined the grams per day of flower consumed and converted that to the average mg THC ingested per day.

However, for participants who consumed both flower and any other cannabis products, we calculated the average mg of THC ingested as follows. For flower, we did the same as for the exclusive flower users and calculated the daily mg of THC from the grams per day. For any of the other products, we used the conversion in table 1 to convert their daily dose captured by the TLFB into mg of THC. We then averaged the amount to get the average daily mg of THC ingested.

## 3. Statistical Analysis

Statistical analyses were conducted in R version 4.3.1. A longitudinal mixed-effects model was fitted using the lmerTest package in R^19^ in order to assess the association between the outcome and each of the covariate variables.

## Results

### 4.1 Characterization of Study Participants

The study population was majority cis-gendered male (59.7%), older (mean age 48.5) and non-Hispanic black (67.2%) PWH. The social-demographic data for this underrepresented population are presented in Table 2.

**Table 2.**
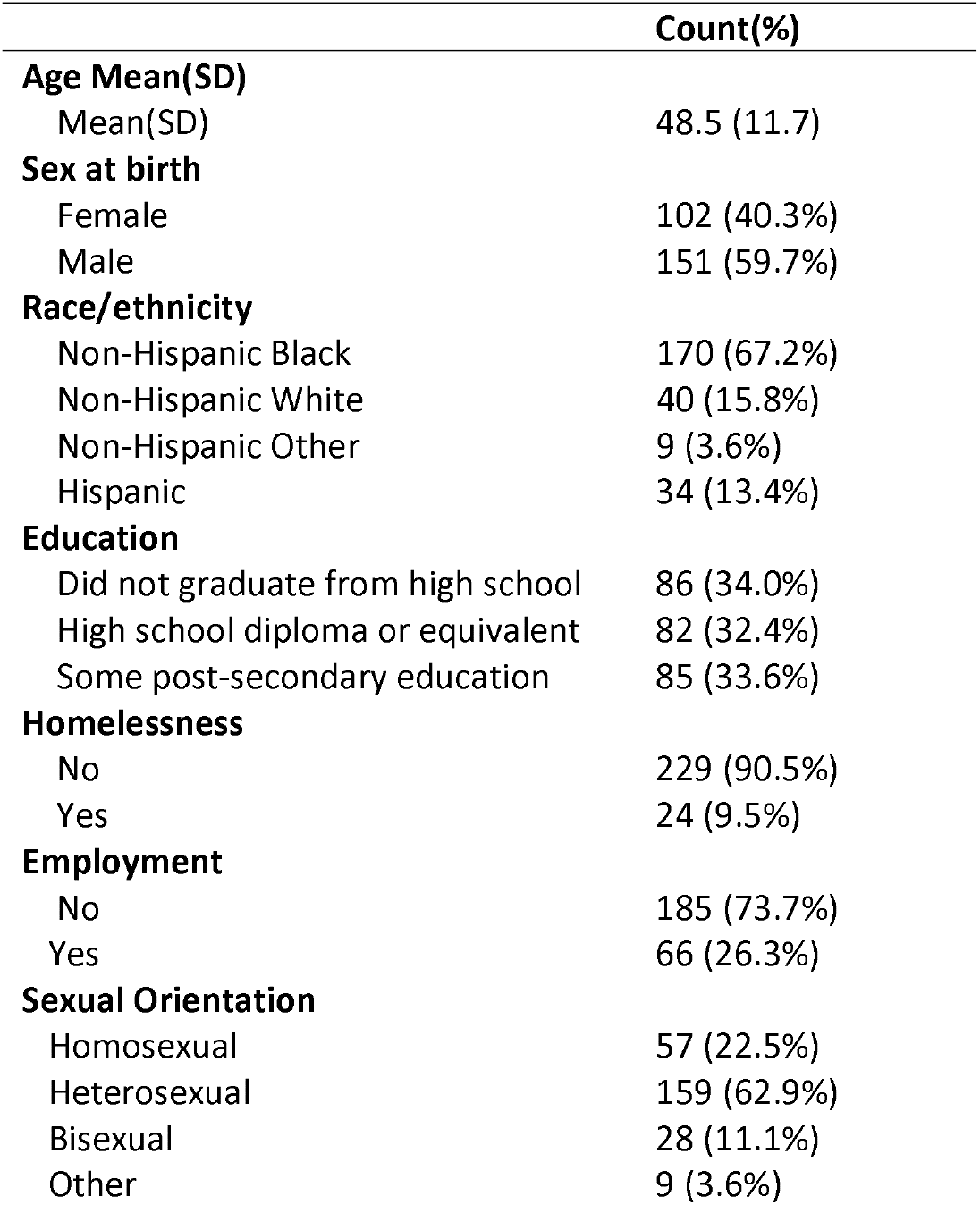
Sample socio-demographics.

|  | Count(%) |
| --- | --- |
| **Age Mean(SD)** |  |
| Mean(SD) | 48.5 (11.7) |
| **Sex at birth** |  |
| Female | 102 (40.3%) |
| Male | 151 (59.7%) |
| **Race/ethnicity** |  |
| Non-Hispanic Black | 170 (67.2%) |
| Non-Hispanic White | 40 (15.8%) |
| Non-Hispanic Other | 9 (3.6%) |
| Hispanic | 34 (13.4%) |
| **Education** |  |
| Did not graduate from high school | 86 (34.0%) |
| High school diploma or equivalent | 82 (32.4%) |
| Some post-secondary education | 85 (33.6%) |
| **Homelessness** |  |
| No | 229 (90.5%) |
| Yes | 24 (9.5%) |
| **Employment** |  |
| No | 185 (73.7%) |
| Yes | 66 (26.3%) |
| **Sexual Orientation** |  |
| Homosexual | 57 (22.5%) |
| Heterosexual | 159 (62.9%) |
| Bisexual | 28 (11.1%) |
| Other | 9 (3.6%) |

### 4.2 Association of average daily mg of THC for exclusively flower users

For the overall group, a one unit increase in the average mg of THC ingested results in a 0.67 increase in the urine THC-COOH biomarker lab result. While for males, we see that a one unit increase results in a 0.57 increase and for women, the same one unit increase results in a 0.88 increase in their lab result. See table 3 for estimates, confidence intervals and p-values. All results were significant at 0.05.

**Table 3.**
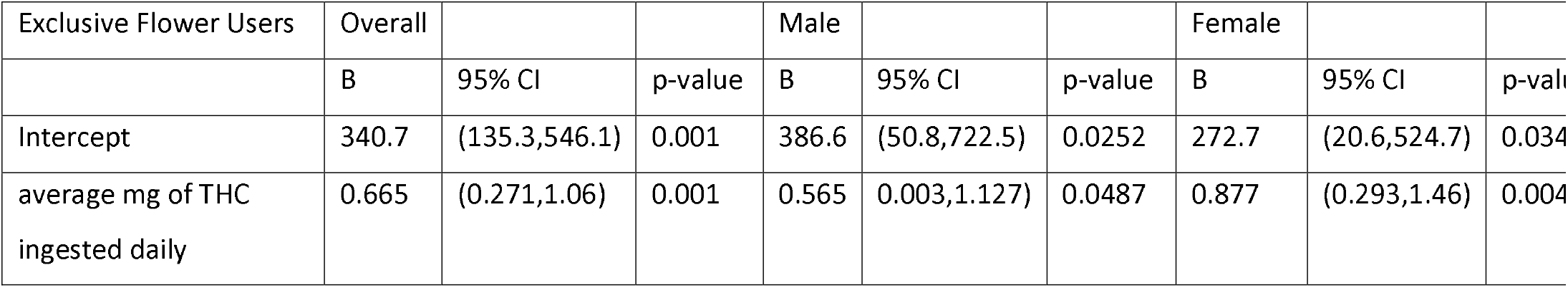

| Exclusive Flower Users | Overall |  |  | Male |  |  | Female |  |  |
| --- | --- | --- | --- | --- | --- | --- | --- | --- | --- |
|  | B | 95% CI | p-value | B | 95% CI | p-value | B | 95% CI | p-value |
| Intercept | 340.7 | (135.3,546.1) | 0.001 | 386.6 | (50.8,722.5) | 0.0252 | 272.7 | (20.6,524.7) | 0.034 |
| average mg of THC ingested daily | 0.665 | (0.271,1.06) | 0.001 | 0.565 | (0.003,1.127) | 0.0487 | 0.877 | (0.293,1.46) | 0.004 |

### 4.3 Association of average daily mg of THC marijuana users of all products

For the overall group, a one unit increase in the average mg of THC ingested daily results in a 0.59 increase in the urine THC-COOH biomarker lab result. Significant at the 0.05 level. However, the results when stratified for males and females were not significant at the 0.05 level. See table 4 for estimates, confidence intervals and p-values.

**Table 4.**

| All product users | Overall |  |  | Male |  |  | Female |  |  |
| --- | --- | --- | --- | --- | --- | --- | --- | --- | --- |
|  | B | 95% CI | p-value | B | 95% CI | p-value | B | 95% CI | p-value |
| Intercept | 457.9 | (201.4,714.4) | <0.001 | 443.5 | (125.8,761.3) | 0.007 | 472.2 | (47.0,897.4) | 0.0301 |
| average mg of THC ingested daily | 0.592 | (0.079,1.10) | 0.024 | 0.526 | (-0.007,1.06) | 0.053 | 0.641 | (-0.371,1.65) | 0.21 |

## 4. Discussion

We found an association between the average mg of THC ingested daily and the THC-COOH urine biomarkers for the both for the overall population and when stratified by gender. The fixed effect was stronger in females than males. This is perhaps due to the lower body mass of women on average in relation to men or perhaps the increased BMI of women to men.

However, when we examine the association between average mg of THC ingested daily and the THC-COOH urine biomarkers for users of all cannabis products, flower included, we only find an association in the overall population and not when stratified by gender. This suggests two limitations in the analysis. First, we are limited by the number of non-flower using participants. Of the 81 marijuana users selected to provide urine samples, only 19 participants were non-flower users, 12 of which were males and 14 of which were females. Second, the way we calculated the THC from products other than flower may not be precise enough or that there is just too much noise in the data. Since the majority of our users obtained their cannabis products from outside the medical marijuana system it is very difficult to know just how much THC was in each dose of the products consumed. We assumed that edible gummies were 10 mg of THC per dose but they range from 10 – 20 mg when purchased from a dispensary and it is common for users to split them. It is also difficult to estimate the amount of THC in homemade chocolate or butter edibles, so we attempted to estimate the amount as 20 mg per dose. And for vapes, we assumed 4 mg per inhalation if the number of puffs were recorded in the TLFB or 10 mg per day but this may not have been accurate enough to capture the signal in the data. See table 1 for details.

Overall, we believe that our timeline follow back successfully captured the amount of flower consumed for the exclusive flower users, but that more research is needed when trying to quantify the THC from products other than flower.

## Data Availability

All data produced in the present study are available upon reasonable request to the authors

